# Side effect profile and comparative tolerability of newer generation antidepressants in the acute treatment of major depressive disorder in children and adolescents: protocol for a systematic review and network meta-analysis

**DOI:** 10.1101/2025.04.02.25324485

**Authors:** Cagdas Türkmen, Seda Sacu, Yuki Furukawa, Angharad N. de Cates, Robert A. Schoevers, Jeanine Kamphuis, Astrid Chevance, John R. Weisz, Graham J. Emslie, Jeffrey R. Strawn, Sarah E. Hetrick, Orestis Efthimiou, Georgia Salanti, Jens H. van Dalfsen, Toshi A. Furukawa, Andrea Cipriani

**Author notes:** **Corresponding author:** Cagdas Türkmen, Central Institute of Mental Health, Department of Addictive Behaviour and Addiction Medicine J5, 68159 Mannheim, Germany. **Authors’ contributions:** CT, guarantor of the review, conceived the study and drafted the protocol. All other authors revised and edited the first draft of the protocol. All authors approved the final version of the protocol to be published. **Patient and public involvement:** Patients or the public were not involved in the design, or conduct, or reporting, or dissemination plans of our research.

## Abstract

**Introduction:** Major depressive disorder (MDD) is among the most common psychiatric disorders in children and adolescents. While previous meta-analyses have synthesised evidence on the efficacy and acceptability of newer-generation antidepressants in this population, specific adverse events (AEs) remain poorly characterised. This is of high clinical importance, as AEs are burdensome for patients, can reduce treatment adherence and lead to discontinuation. Here, we present a protocol for a network meta-analysis designed to evaluate the specific AE profile and comparative tolerability of newer-generation antidepressants in children and adolescents with MDD.

**Methods and analysis:** The planned study will include double-blind randomised controlled trials that compared one active drug with another and/or placebo for the acute treatment of MDD in children and adolescents. The following antidepressants will be considered: agomelatine, bupropion, citalopram, desvenlafaxine, duloxetine, escitalopram, fluoxetine, fluvoxamine, milnacipran, mirtazapine, paroxetine, reboxetine, sertraline, venlafaxine, alaproclate, vilazodone, vortioxetine, levomilnacipran, and edivoxetine. The primary outcomes will include the number of patients experiencing at least one AE, specific non-serious AEs, serious AEs (e.g., suicidal ideation), and AEs leading to treatment discontinuation. Published and unpublished studies will be retrieved through a systematic search in the following databases: PubMed, Embase, Cochrane Library (including the Cochrane Central Register of Controlled Trials), Web of Science Core Collection, PsycInfo and regulatory agencies’ registries. Study selection and data extraction will be performed independently by two reviewers. For each outcome, a network meta-analysis will be performed to synthesise all evidence. Consistency will be assessed through local and global methods, and the confidence in the evidence will be evaluated using the Confidence in Network Meta-Analysis (CINeMA) web application. All analyses will be conducted in the R software.

**Ethics and dissemination:** The planned review does not require ethical approval. The findings will be published in a peer-reviewed journal and may be presented at international conferences.

**PROSPERO registration number:** CRD420251011399.

**Strengths and limitations of this study:**

- This will be the most comprehensive network meta-analysis on the safety of newer generation antidepressants in children and adolescents with major depression, incorporating both published and unpublished data.
- Consistency will be assessed using both local and global methods, and the robustness of the results will be examined through network meta-regression.
- Antidepressant trials, particularly placebo-controlled trials, present a high risk of selection bias regarding adverse events.
- Study limitations will be addressed with the Cochrane Risk of Bias 2 tool, and the confidence in the evidence for network estimates of the main outcomes will be assessed with the GRADE framework.

## INTRODUCTION

Major depressive disorder (MDD) represents a common public health issue, with an estimated point prevalence of 8% among youth aged 10 to 19 years.^1^ An early onset of depression has been associated with multiple adverse outcomes in adulthood, including a threefold risk of depression, unemployment, increased levels of anxiety and illicit drug disorders, lower educational attainment, as well as poorer physical health and social functioning.^2-4^ Depressive disorders are among the leading contributors to the global burden of disease among young people aged 10 to 24 years, highlighting the need for accessible, effective and safe treatments.^5^

Newer generation antidepressants, including selective serotonin reuptake inhibitors (SSRIs) and serotonin-norepinephrine reuptake inhibitors (SNRIs), are the primary pharmacologic treatment options. Current guidelines recommend SSRIs, either alone or in combination with cognitive behavioural therapy or interpersonal therapy, as the first-line treatment for moderate-to-severe MDD in youth.^6 7^ While previous network meta-analyses (NMAs) of randomised controlled trials suggest a modest benefit of SSRIs and SNRIs, clinicians must individually weigh both the benefits and risks of the respective antidepressant treatment.^8-10^ Both SSRIs and SNRIs are associated with acute and late-emergent adverse events (AEs) in youth, which can reduce treatment adherence and increase discontinuation rates, ultimately undermining treatment success.^8-11^

While prior NMAs have identified variability in acceptability (all-cause discontinuation or discontinuation due to AEs) among antidepressants,^8 9^ specific AE profiles in youth with MDD remain poorly characterised. Among the AEs, most research has focused specifically on suicidal behaviour or ideation,^8 10^ following the U.S. Food and Drug Administration’s boxed warning on antidepressant-related suicide risk in young people, which remains a matter of controversy.^12^ A recent meta-analysis expanded the evidence base by examining insomnia as an AE in youth with MDD.^13^ This study found that SSRIs and SNRIs are associated with a mostly increased risk of treatment-emergent insomnia during acute treatment, with no difference between SSRIs and SNRIs.^13^ However, there was substantial variability regarding this risk among individual antidepressants.^13^ Expanding the line of research on specific AEs could provide a more precise understanding of the AE profile of individual antidepressants, which may improve clinical decision-making and contribute to treatment guidelines.

The aim of the planned study – Project **SOTERIA** (**S**afety **O**utcomes and **T**olerability: **E**valuating and **R**evisiting the **I**nitiation of **A**ntidepressants) – is to evaluate the specific AE profile and comparative tolerability of newer generation antidepressants in children and adolescents with MDD. It will complement an ongoing NMA that evaluates the AE profile and comparative tolerability of 21 antidepressants in adults with MDD.^14^

## METHODS AND ANALYSIS

The information outlined in this protocol is reported in accordance with the guidelines of the Preferred Reporting Items for Systematic Review and Meta-Analysis Protocols (PRISMA-P).^15 16^ The completed PRISMA-P 2015 Checklist is available in **Supplement 1**. The planned NMA will use the same eligibility criteria as a recent meta-analysis, except that it will also include head-to-head trials.^13^ The protocol was prospectively registered on PROSPERO (CRD420251011399).

### Types of studies

We will include randomised controlled trials (RCTs) comparing the antidepressants of interest (see **Types of interventions**) with one another and/or placebo in the acute phase of antidepressant treatment (min. 6 weeks and max. 12 weeks) in a double-blind, parallel-group fashion. Only monotherapy trials will be included (i.e., those in which the antidepressant is used an add-on treatment or where additional interventions are evaluated will be excluded). Quasi-randomised trials will be excluded, while cluster randomised trials will be included. Longer-term RCTs will be considered, provided that they report data for the acute treatment period (initial 6 to 12 weeks). Both fixed- and flexible-dose designs will be included. RCTs permitting the use of any rescue / pro re nata (PRN) medications will be included.

### Types of participants

The population of interest comprises children and adolescents (≤ 18 years of age), of any gender, with a primary diagnosis of MDD. RCTs employing standard operationalised diagnostic criteria such as the *Diagnostic and Statistical Manual of Mental Disorders (DSM)-III, DSM-IV, DSM-5*, or the International Classification of Diseases (ICD)-10 and ICD-11 will be included. Secondary psychiatric comorbidities will not be grounds for exclusion. RCTs in which MDD is not the primary focus or is a comorbidity of a primary psychiatric condition (e.g., eating disorder or attention-deficit/hyperactivity disorder) will be excluded.

### Types of interventions

The planned study will evaluate the following newer-generation antidepressants: agomelatine, bupropion, citalopram, desvenlafaxine, duloxetine, escitalopram, fluoxetine, fluvoxamine, milnacipran, mirtazapine, paroxetine, reboxetine, sertraline, venlafaxine, alaproclate, vilazodone, vortioxetine, levomilnacipran and edivoxetine. Data will be obtained from both head-to-head and placebo-controlled trials, in which antidepressants are administered within their licensed dose range. **Figure 1** illustrates the network of all possible pairwise comparisons between the interventions. We anticipate that any patient meeting the inclusion criteria could, in principle, be randomly assigned to any of the interventions within the synthesis comparator set (transitivity assumption).

**Figure 1.**
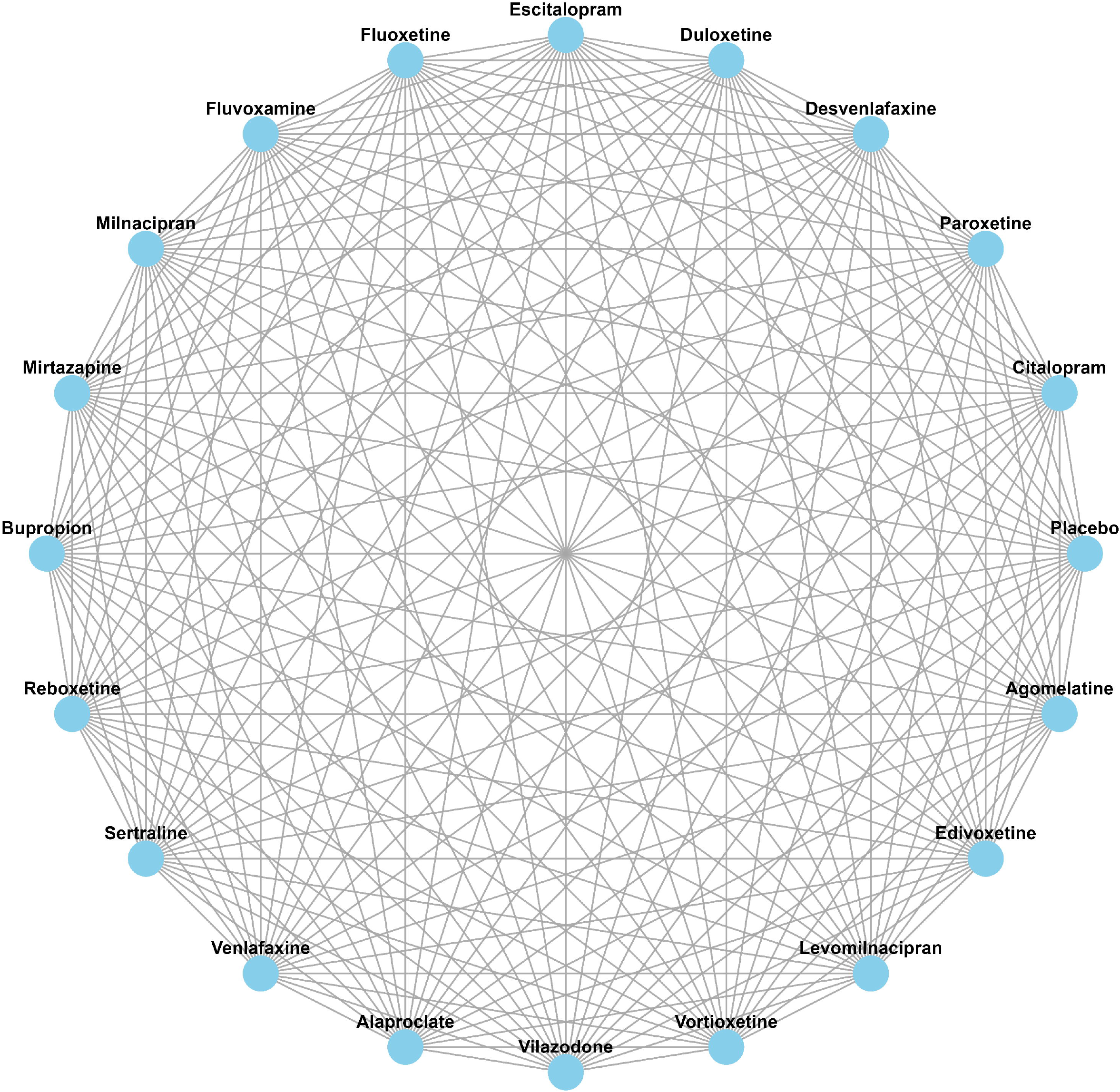
Network of possible pairwise comparisons between the eligible interventions.

### Outcomes and categorisation of AEs

The tolerability of the antidepressants will be evaluated using the following outcomes:

1. Total number of patients experiencing serious AEs (e.g., suicidal ideation; further details below).
2. Total number of patients experiencing AEs leading to treatment discontinuation.
3. Total number of patients experiencing one specific AE (e.g., somnolence).
4. Total number of patients experiencing at least one AE.

Two independent reviewers will extract all AEs reported in the trials, ensuring that no events are double-counted. The reviewers will then use the preferred terms from the Medical Dictionary for

Regulatory Activities (MedDRA) (https://www.meddra.org/) to categorise each AE. Details regarding MedDRA are described elsewhere.^14^ If different MedDRA terms are used to describe similar AEs, these will be merged into broader categories based on clinical judgment and validated by another member of the review team. Any disagreements will be resolved through consensus within the review team.

As a definition for serious AEs, we will use the classification used by the U.S. Food and Drug Administration (FDA) (https://www.fda.gov/):

▸Resulted in death.
▸Life-threatening.
▸Hospitalisation (initial or prolonged).
▸Disability or permanent damage.
▸Congenital anomaly/birth defect.
▸Required intervention to prevent permanent impairment or damage (devices).
▸Other serious (important medical events), e.g., seizures that did not result in hospitalisation.

All serious AEs will be included in the meta-analysis. The following variables have been selected as the most important outcomes to be evaluated, given their inclusion in FDA’s boxed warning^12^ and/or their high clinical relevance, and will be subject to risk of bias and certainty of evidence assessments:

1. suicidal ideation,
2. suicidal behaviour, and
3. discontinuation due to AEs.

To determine which specific non-serious AEs should be included in the statistical analyses, we will incorporate findings on core AE outcomes from the ongoing International Network for Research Outcomes in Adolescent Depression Studies (IN-ROADS) project (https://www.in-roads.org/).^17^ However, if the findings regarding core AE outcomes are not available at the time of analysis and/or if IN-ROADS project members are not able to provide such information, we will use an alternative approach that considers both the frequency of AEs in our meta-analyses and the perceived importance of AEs based on the findings of a recent international study that aimed to identify the most important non-serious AEs of antidepressants according to adult patients’ rankings.^18^ We will select up to 10 AEs which are both the most common in our meta-analyses and ranked as the most important AEs by adult patients.^18^ However, the review team will evaluate and exclude AEs considered to be less relevant to children and adolescents (e.g., sexual dysfunction).

### Search strategy and study selection

The planned study will adopt the same search strategy as in a recent meta-analysis,^13^ and perform an update of the search. As the previous study focused on insomnia as an AE, sleep-related search terms will be removed from the search strings. Additionally, RCTs that were previously excluded solely due to the absence of reported data on insomnia as an AE or the lack of a placebo group will be reassessed for eligibility. The following databases will be systematically searched to identify both published and unpublished RCTs from 31/08/2023 (the search end date in the previous meta-analysis^13^) through the date of the search: PubMed, Embase, Cochrane Library (including the Cochrane Central Register of Controlled Trials for unpublished RCTs), Web of Science Core Collection and PsycInfo. No language restrictions will be applied. Regulatory agencies’ registries (e.g., U.S. Food and Drug Administration) will also be searched for relevant studies and/or data. The search will be supplemented by screening the reference lists of newly published studies that meet the eligibility criteria since the recent meta-analysis.^13^ The search strings are provided in **Supplement 2**.

Two reviewers will independently screen the titles and abstracts of potentially relevant studies identified in the previous meta-analysis (described in the Supplementary Materials of the previous meta-analysis^13^) and newly published studies since the date of the last search. Full-text reports of potentially relevant studies will be retrieved and assessed for eligibility. Disagreements regarding study selection will be resolved by consensus or, if necessary, with the involvement of another member of the review team. Inter-rater agreement will be reported in terms of percentage agreement and Cohen’s kappa.^19^

### Record management

Records that have been identified through the systematic search will first be imported into EndNote (Clarivate, Version 21, 2023)^20^ and deduplicated using the software’s built-in functions. After deduplication, the records will be exported from EndNote to an Excel spreadsheet for formal screening. At this stage, records will be coded as either clearly irrelevant (excluded prior to full-text review based on title and/or abstract) or potentially relevant (to be assessed at the full-text review stage). Following full-text review, potentially relevant records will be coded as either included or excluded, with reasons for exclusion documented for each record.

### Data extraction

Two reviewers will independently extract data from all included RCTs for the following clinical and methodological variables: authors, year of publication, diagnostic criteria, name of antidepressant and dose range, treatment duration, number of participants, age range (and mean), proportion of young people who are females, country of recruitment site, patient setting (e.g., outpatient vs inpatient), race/ethnicity (when available) and funder. This information will be summarised narratively in the text and/or presented in tabular form.

Categorical data (yes/no) for the outcomes of interest (AEs) will be extracted using a predefined structured extraction sheet. Efforts will be made to obtain unpublished data on AEs from trial registries and study summaries from drug company websites, as there is strong evidence indicating that a large amount of information on AE remains unpublished and that both the number and scope of AEs are greater in unpublished versions compared to published versions of the same study.^13 21 22^ Pharmaceutical companies will be contacted to request additional data.

As discrepancies in AE data (e.g., between publications and FDA reports) may occur, we will prioritise data sources based on the following hierarchy, where applicable:

1. FDA / regulatory agency reports
2. Reanalyses by independent groups (e.g., reanalysis of Study 329^22^)
3. Company trial documents
4. Trial registry entries
5. Peer-reviewed publications

### Statistical synthesis of data

The planned NMA will adopt the same statistical analysis approach as an ongoing NMA that evaluates the AE profile and comparative tolerability of antidepressants in adults with MDD.^14^ If a quantitative synthesis is deemed inappropriate, we will provide a systematic narrative synthesis summarising study characteristics and findings in text and/or table(s). The narrative synthesis will evaluate both within-study and between-study results.

### Pairwise meta-analyses

For pairwise comparisons informed by ≥ 10 RCTs, a random-effects meta-analysis model will be used to obtain odds ratios (ORs) and 95% confidence intervals (CIs), based on the assumption that treatment effects are similar but not identical across study settings.^23^ Based on previous research, we expect that some AEs, particularly suicide-related outcomes, will be rare (i.e., low counts) in some studies.^10 13^ For rare AEs, the Mantel-Haenszel method will be used, which avoids continuity corrections that might bias results.^24 25^ We will compare the results of the inverse variance model (which assumes a common treatment effect) and the Mantel-Haenszel method. If there are considerable discrepancies, only the Mantel-Haenszel method will be used. Forest plots will be presented for all pairwise meta-analyses, with a 0.5 continuity correction for studies with zero events in one treatment arm. We will visually inspect the forest plots for any heterogeneity and report the I^2^ statistic, along with its 95% CIs, for all analyses. As an additional way of assessing heterogeneity, we will present prediction intervals.^26 27^

### Assessment of the transitivity assumption of NMA

Transitivity is a key underlying assumption of NMA.^28 29^ To assess its validity, we will examine whether study-level characteristics that could affect relative treatment effects, such as age, sex, depressive severity at baseline assessment, and dosing schedule. If significant discrepancies are found, we will restrict our NMA to studies with similar distributions. Furthermore, to ensure transitivity in our network, RCTs in which MDD is not the primary focus or is a comorbidity of a primary psychiatric condition (e.g., eating disorder or attention-deficit/hyperactivity disorder) will be excluded.

### Network meta-analyses

If the transitivity assumption holds, we will synthesise the evidence using NMA.^29^ For outcomes that are not rare, a random-effects frequentist NMA model will be used, which assumes a common heterogeneity parameter across all comparisons.^30^ Results will be presented in a “league table” including estimated treatment effects and corresponding 95% CIs. To assess heterogeneity, we will compare the estimated value for the heterogeneity standard deviation with the corresponding empirical distributions^31^ and report the prediction interval for each outcome.^26 27^

For rare outcomes (i.e., with zero events in a treatment arm), we will use a fixed-effects Mantel-Haenszel NMA model without continuity correction^32^ and compare the results with the inverse variance NMA model. If results are consistent, we will use the random-effects NMA model^30^; in the case of discrepancies between the approaches, we will only present results from the Mantel-Haenszel NMA approach. We will additionally perform sensitivity analyses using an NMA model with a non-central hypergeometric (NCH) likelihood.^33^ In the case of very rare outcomes, we expect that the network might become disconnected. In that case, we will perform NMAs in each of the corresponding subnetworks that include enough data to be meaningfully synthesised.

### Assessment of inconsistency

Inconsistency refers to statistical disagreement between different sources of evidence in a network, which can challenge the validity of the transitivity assumption in NMA.^29^ To assess inconsistency, we will use two methods, namely a global method (the design-by-treatment test)^34^ and a local method (“Separate Indirect from Direct Design Evidence”; SIDDE).^32^ The global method tests the null hypothesis of overall consistency in the network, while the local method compares direct and indirect evidence for each treatment comparison to detect “hot spots” of inconsistency in the network.

If inconsistency is detected, we will initially check for data extraction errors and reassess the plausibility of transitivity, particularly in the presence of hot spots. If the cause of inconsistency cannot be identified, we will interpret the NMA results with caution. It is important to note that tests for inconsistency may have low power in detecting violations of the transitivity assumption, particularly when outcomes are rare. Therefore, we will carefully assess the transitivity assumption, even in the absence of evidence for inconsistency.

### Exploring heterogeneity and inconsistency and sensitivity analyses

Given the variety of study settings we intend to include, we anticipate at least a small degree of heterogeneity and inconsistency. For all outcomes, we will examine whether treatment effects remain consistent through subgroup analyses and network meta-regression if enough data become available, considering the following factors: (1) age (children vs adolescents), (2) sex, (3) depressive severity at baseline assessment, and (4) dosing schedule. If few studies are available per comparison, we will instead group all antidepressants together and conduct pairwise meta-regressions comparing antidepressants with placebo.

To assess the robustness of our conclusions, we will conduct sensitivity analyses by evaluating (1) only studies with unpublished data (excluding those with only published data) and (2) only studies without a high risk of bias for the respective outcome.

### Assessing small study effects, publication bias and reporting bias

Antidepressant trials, particularly placebo-controlled trials, present a high risk of reporting bias regarding AEs.^35-37^ Provided that ≥ 10 studies are included,^38^ the presence of potential publication bias or ‘small-study effects’ will be assessed by examining asymmetry in a contour-enhanced funnel plot where all drugs are compared against the placebo comparison.^39^ The Harbord test will be used to formally test for asymmetries in the funnel plots.

### Model implementation

All models will be fitted in the R software using the meta^40^ package for the pairwise meta-analysis models and the netmeta^41^ package for the frequentist NMAs

### Risk of bias

Two reviewers will independently assess the risk of bias for the three most important outcomes (suicidal ideation, suicidal behaviour and discontinuation due to AEs) in each study using the Cochrane Risk of Bias 2 (RoB 2) tool.^42^ The following domains will be assessed:

1. bias arising from the randomisation process,
2. bias due to deviations from the intended interventions,
3. bias due to missing outcome data,
4. bias in the measurement of the outcome, and
5. bias in the selection of the reported result.

Each domain will be rated as “low risk of bias”, “some concerns”, or “high risk of bias”. The overall risk of bias for each outcome will be determined by the least favorable rating among the domains. The assessments will be managed using the RoB 2 Excel tool. Disagreements regarding the RoB 2 ratings will be resolved by consensus or, if necessary, with the involvement of another member of the review team. Inter-rater agreement will be reported in terms of percentage agreement and Cohen’s kappa.^19^

Reporting bias (bias due to missing evidence), which is a particularly important issue in antidepressant trials,^13 43 44^ will be evaluated for the three most important outcomes (suicidal ideation, suicidal behaviour and discontinuation due to AEs) using the Risk Of Bias due to Missing Evidence in Network meta-analysis (ROB-MEN) web application.^45 46^

### Certainty of evidence of NMA

Two reviewers will independently assess the certainty of evidence for the three most important outcomes (suicidal ideation, suicidal behaviour and discontinuation due to AEs) using the framework described in Salanti et al’s study.^47^ We will apply a margin of equivalence of 0.95–1.05 for the ORs of the three most important outcomes (suicidal ideation, suicidal behaviour and discontinuation due to AEs).^48^ This narrow margin was selected due to the clinical importance of even small differences in these outcomes. The Confidence in Network Meta-Analysis (CINeMA)^48 49^ web application will be used to complete the assessments. The certainty in the body of evidence will be rated as high, moderate, low or very low. Justifications will be provided for decisions to downgrade or upgrade the certainty of the evidence, as well as for the importance rating of each outcome.

## DISCUSSION

AEs that emerge during the acute phase of antidepressant treatment pose a significant clinical challenge, potentially leading to reduced treatment adherence and higher discontinuation rates.^8-11^ While prior studies have primarily assessed serious AEs, specifically suicidal behaviour or ideation,^8 10 50 51^ more precise understanding of other (non-serious) AEs is significantly lacking. Such an understanding may help clinicians better balance the risks and benefits of antidepressant treatments, ultimately improving patient care. The planned study aims to provide a comprehensive evaluation of AEs associated with newer-generation antidepressants, which may help tailor antidepressant treatment to the specific needs of children and adolescents with MDD. By refining risk assessment and optimising prescribing, these findings have the potential to improve clinical decision-making and refine clinical guidelines for the treatment of children and adolescents with MDD.

## Supporting information

Supplement 1. PRISMA-P 2015 Checklist

Supplement 2. Search strategy

## Data Availability

Not applicable, as this is a protocol for a systematic review.

## Acknowledgements

We thank the librarian Dipl.-Bibl. Volker Braun from the Library of the Medical Faculty Mannheim, University of Heidelberg, for his assistance in developing the systematic search strategy and managing the records. We also extend our gratitude to the Central Institute of Mental Health in Mannheim, Germany, for supporting this work through institutional open access funding.

ACi is supported by the National Institute for Health Research (NIHR) Oxford Cognitive Health Clinical Research Facility, by an NIHR Research Professorship (grant RP-2017-08-ST2-006), by the NIHR Oxford and Thames Valley Applied Research Collaboration, by the NIHR Oxford Health Biomedical Research Centre (grant NIHR203316) and by the Wellcome Trust (GALENOS Project). AdeC is supported by an NIHR Academic Clinical Lectureship and by the NIHR Oxford Health Biomedical Research Centre (grant NIHR203316). The views expressed are those of the authors and not necessarily those of the UK National Health Service, the NIHR, or the UK Department of Health and Social Care.

