## Supplement 2. Search strategy for "Side effect profile and comparative tolerability of newer generation antidepressants in the acute treatment of major depressive disorder in children and adolescents: protocol for a systematic review and network meta-analysis"

**We will use the same strategy used in Türkmen et al. (2025)^1^ with the exception that sleep-related search terms will be removed (indicated by strikethrough in the search syntaxes below), and the search will be updated accordingly.**

1. Türkmen C, Machunze N, Lee AM, et al. Systematic Review and Meta-Analysis: The Association Between Newer Generation Antidepressants and Insomnia in Children and Adolescents With Major Depressive Disorder. *Journal of the American Academy of Child & Adolescent Psychiatry* 2025 doi: 10.1016/j.jaac.2025.01.006

### 1.1 PubMed

**08/31/2023, 1011 records**

(

"Antidepressive Agents, Second-Generation"[Mesh] OR

"Antidepressive Agents, Second-Generation"[Pharmacological Action] OR

"Second Generation Antidepress*"[tiab] OR

"Selective Serotonin Reuptake Inhibitors"[Mesh] OR

"Serotonin and Noradrenaline Reuptake Inhibitors"[Mesh] OR

"Serotonin Uptake Inhibitor"[tiab:~2] OR

"Serotonin Uptake Inhibitors"[tiab:~2] OR

"Serotonin Reuptake Inhibitor"[tiab:~2] OR

"Serotonin Reuptake Inhibitors"[tiab:~2] OR

SSRI*[tiab] OR

SNRI*[tiab] OR

agomelatine[tw] OR

bupropion[tw] OR

citalopram[tw] OR

desvenlafaxine[tw] OR

duloxetine[tw] OR

escitalopram[tw] OR

fluoxetine[tw] OR

fluvoxamine[tw] OR

milnacipran[tw] OR

mirtazapine[tw] OR

paroxetine[tw] OR

reboxetine[tw] OR

sertraline[tw] OR

venlafaxine[tw] OR

alaproclate[tw] OR

vilazodone[tw] OR

vortioxetine[tw] OR

levomilnacipran[tw] OR

edivoxetine[tw]

)

AND

(

"Depressive Disorder, Major"[Mesh] OR

"major depressive disorder*"[tiab] OR

"major depression"[tiab] ~~OR~~

~~"Sleep Initiation and Maintenance Disorders"[Mesh] OR~~

~~Insomnia*[tiab] OR~~

~~Sleep*[tw]~~

)

AND

(

(adolescent[mh] OR child[mh] OR pediatrics[mh] OR

minors* [tiab] OR boy [tiab] OR boys [tiab] OR boyhood [tiab] OR girl* [tiab] OR kid [tiab] OR kids [tiab] OR child* [tiab] OR adolescen* [tiab] OR juvenil* [tiab] OR youth* [tiab] OR teen* [tiab] OR tween* [tiab] OR prepubescen* [tiab] OR pubescen* [tiab] OR

"under age*" [tiab] OR underage* [tiab] OR pediatric* [tiab] OR paediatric* [tiab] OR school* [tiab] OR "young people*" [tiab] OR "young person*" [tiab])

NOT (animals[mh] NOT humans[mh])

NOT (adult[mh] NOT (adolescent[mh] OR child[mh] OR infant[mh]))

)

AND

(

"Controlled Clinical Trial"[pt] OR "clinical trials as topic"[mesh] OR "clinical trial*"[tiab] OR random*[tiab] OR placebo[tiab] OR trial[ti]

)

NOT (review[pt] OR meta-analysis[pt] OR "Systematic Review" [pt])

### 1.2 Embase

**08/31/2023, 1104 records**

(

'antidepressant agent'/de OR

"Second Generation Antidepress*":ti,ab,kw OR

'serotonin uptake inhibitor'/exp OR

'serotonin noradrenalin reuptake inhibitor'/exp OR

(Serotonin NEXT/3 "Uptake Inhibitor*"):ti,ab,kw OR

(Serotonin NEXT/3 "Reuptake Inhibitor*"):ti,ab,kw OR

SSRI*:ti,ab,kw OR

SNRI*:ti,ab,kw OR

agomelatine OR

bupropion OR

citalopram OR

desvenlafaxine OR

duloxetine OR

escitalopram OR

fluoxetine OR

fluvoxamine OR

milnacipran OR

mirtazapine OR

paroxetine OR

reboxetine OR

sertraline OR

venlafaxine OR

alaproclate OR

vilazodone OR

vortioxetine OR

levomilnacipran OR

edivoxetine

)

AND

(

'major depression'/exp OR

"major depressive disorder*":ti,ab,kw OR

"major depression":ti,ab,kw ~~OR~~

~~'insomnia'/exp OR~~

~~Insomnia*:ti,ab,kw OR~~

~~Sleep*:ti,ab,kw,de~~

)

AND

(

('juvenile'/exp OR 'pediatrics'/exp OR

minors*:ti,ab,kw OR boy:ti,ab,kw OR boys:ti,ab,kw OR boyhood:ti,ab,kw OR girl*:ti,ab,kw OR kid:ti,ab,kw OR kids:ti,ab,kw OR child*:ti,ab,kw OR adolescen*:ti,ab,kw OR juvenil*:ti,ab,kw OR youth*:ti,ab,kw OR teen*:ti,ab,kw OR tween*:ti,ab,kw OR prepubescen*:ti,ab,kw OR pubescen*:ti,ab,kw OR

"under age*":ti,ab,kw OR underage*:ti,ab,kw OR pediatric*:ti,ab,kw OR paediatric*:ti,ab,kw OR school*:ti,ab,kw OR "young people*":ti,ab,kw OR "young person*":ti,ab,kw)

NOT ([animals]/lim NOT [humans]/lim)

NOT ('adult'/exp NOT 'juvenile'/exp)

)

AND

(

'controlled clinical trial'/exp OR 'clinical trial (topic)'/exp OR "clinical trial*":ti,ab,kw OR random*:ti,ab,kw OR placebo:ti,ab,kw OR trial:ti

)

NOT ('review'/exp OR 'meta analysis'/exp)

### 1.3 Cochrane Library

**08/31/2023, 1207 records**

(

Second NEXT Generation NEXT Antidepress* OR

Serotonin NEAR/3 *uptake NEXT Inhibitor* OR

SSRI* OR

SNRI* OR

agomelatine OR

bupropion OR

citalopram OR

desvenlafaxine OR

duloxetine OR

escitalopram OR

fluoxetine OR

fluvoxamine OR

milnacipran OR

mirtazapine OR

paroxetine OR

reboxetine OR

sertraline OR

venlafaxine OR

alaproclate OR

vilazodone OR

vortioxetine OR

levomilnacipran OR

edivoxetine

):ti,ab,kw

AND

(

Major NEXT depressive NEXT disorder* OR

"major depression" ~~OR~~

~~Insomnia* OR~~

~~Sleep*~~

):ti,ab,kw

AND

(

minors* OR boy OR boys OR boyhood OR girl* OR kid OR kids OR child* OR adolescen* OR juvenil* OR youth* OR teen* OR tween* OR prepubescen* OR pubescen* OR under NEXT age* OR underage* OR pediatric* OR paediatric* OR school* OR young NEXT people* OR young NEXT person*

):ti,ab,kw

### 1.4 Web of Science Core Collection

**08/31/2023, 719 records**

TS=(

"Second Generation Antidepress*" OR

Serotonin NEAR/2 "*uptake Inhibitor*" OR

SSRI* OR

SNRI* OR

agomelatine OR

bupropion OR

citalopram OR

desvenlafaxine OR

duloxetine OR

escitalopram OR

fluoxetine OR

fluvoxamine OR

milnacipran OR

mirtazapine OR

paroxetine OR

reboxetine OR

sertraline OR

venlafaxine OR

alaproclate OR

vilazodone OR

vortioxetine OR

levomilnacipran OR

edivoxetine

)

AND

TS=(

"Major depressive disorder*" OR

"major depression" ~~OR~~

~~Insomnia* OR~~

~~Sleep*~~

)

AND

(

TS=("clinical trial*" OR

random* OR

placebo) OR

(TI=trial)

)

AND

TS=(

minors* OR boy OR boys OR boyhood OR girl* OR kid OR kids OR child* OR adolescen* OR juvenil* OR youth* OR teen* OR tween* OR prepubescen* OR pubescen* OR "under age*" OR underage* OR pediatric* OR paediatric* OR school* OR "young people*" OR "young person*"

)

### 1.5 PsycInfo (EBSCO)

**08/31/2023, 375 records**

(

“Second Generation Antidepress*” OR

Serotonin N2 “*uptake Inhibitor*” OR

SSRI* OR

SNRI* OR

agomelatine OR

bupropion OR

citalopram OR

desvenlafaxine OR

duloxetine OR

escitalopram OR

fluoxetine OR

fluvoxamine OR

milnacipran OR

mirtazapine OR

paroxetine OR

reboxetine OR

sertraline OR

venlafaxine OR

alaproclate OR

vilazodone OR

vortioxetine OR

levomilnacipran OR

edivoxetine

)

AND

(

“Major depressive disorder*” OR

“major depression” ~~OR~~

~~Insomnia* OR~~

~~Sleep*~~

)

AND

(

(“clinical trial*” OR

random* OR

placebo) OR

(TI trial)

)

AND

(

AG “childhood (birth-12 yrs)” OR AG “adolescence (13-17 yrs)”

)
